# Evaluating Chatbots in Psychiatry: Rasch-Based Insights into Clinical Knowledge and Reasoning

**DOI:** 10.1101/2024.12.30.24319383

**Authors:** Yu Chang, Si-Sheng Huang, Wen-Yu Hsu, Yi-Chun Liu

**Affiliations:** Department of Psychiatry, Changhua Christian Hospital, Changhua 500, Taiwan; Department of Psychiatry, Changhua Christian Children’s Hospital, Changhua 500, Taiwan; Department of Health Business Administration, Hungkuang University, Taichung City, Taiwan

**Keywords:** chatbot, psychiatry, clinical knowledge, clinical reasoning, Rasch model

## Abstract

Chatbots are increasingly being recognized as valuable tools for clinical support in psychiatry. This study systematically evaluates their strengths and limitations in psychiatric clinical knowledge and reasoning. A total of 27 chatbots, including ChatGPT-o1-preview, were assessed using 160 multiple-choice questions derived from the 2023 and 2024 Taiwan Psychiatry Licensing Examinations. The Rasch model was employed to analyze chatbot performance, supplemented by dimensionality analysis and qualitative assessments of reasoning processes. Among the models, ChatGPT-o1-preview achieved the highest performance, with a JMLE ability score of 2.23, significantly exceeding the passing threshold (p < 0.001). It excelled in diagnostic and treatment reasoning and demonstrated a strong grasp of psychopharmacology concepts. However, limitations were identified in its factual recall, handling of niche topics, and occasional reasoning biases. Building on these findings, we have highlighted key aspects of a potential clinical workflow to guide the practical integration of chatbots into psychiatric practice. While ChatGPT-o1-preview holds significant potential as a clinical decision-support tool, its limitations underscore the necessity of human oversight. Continuous evaluation and domain-specific training are crucial to maximize its utility and ensure safe clinical implementation.

## 1. Introduction

Chatbots, powered by generative artificial intelligence and trained through deep learning algorithms, are designed to engage in natural conversations ^1^. These systems, often referred to as large language models (LLMs), exhibit a remarkable ability to process and respond with contextually relevant information ^1^. Recent advancements in large-scale data training and sophisticated reasoning mechanisms have expanded chatbots’ capabilities from general knowledge dissemination to specialized applications ^2^. In the medical field, research has demonstrated that chatbots can achieve acceptable levels of performance in various professional examinations and assessments ^3-8^.

The growing interest in chatbots for clinical support highlights their potential to enhance healthcare delivery ^9^. Previous studies have established a connection between clinical competence and foundational knowledge ^10^, suggesting that an evaluation of chatbots’ clinical knowledge could provide valuable insights into their practical utility. While previous research has used Rasch analysis to assess chatbots’ knowledge in psychiatry, gaps in understanding remain, particularly in certain specialized areas ^4^. A comprehensive evaluation of chatbots’ strengths and limitations in psychiatric clinical knowledge is still needed.

Measuring psychiatric clinical knowledge poses unique challenges due to the complexity and context-dependent nature of psychiatric assessments. Past research has attempted to evaluate chatbots in this domain ^6, 11, 12^, with one study administering a set of ten multiple-choice questions focused on differential diagnosis to both chatbots and psychiatrists ^6^. However, the study primarily relied on surface-level comparisons of scores and lacked sufficient depth to quantify or qualitatively understand the underlying knowledge structures of the chatbots. This limited scope has left aspects of chatbot capabilities, such as their reasoning processes and domain-specific knowledge gaps, underexplored.

To address these limitations, this study employs Rasch analysis ^13^, a statistical method commonly used in educational and psychological testing to evaluate item difficulty and respondent ability on a unified scale. By applying Rasch analysis to a larger and more diverse set of psychiatry-related multiple-choice questions, we aim to provide a detailed assessment of the strengths and weaknesses of the most advanced chatbots. Additionally, this study explores a broader range of chatbot models to ensure a more comprehensive evaluation, offering deeper insights into their capabilities and limitations across different psychiatric domains.

## 2. Methods

### 2.1. Study Design and Question Selection

To evaluate the clinical knowledge of chatbots, this study utilized multiple-choice questions (MCQs) from the Taiwan Psychiatry Licensing Examination administered by the Taiwanese Society of Psychiatry. To comprehensively evaluate chatbot performance, we included questions related to any clinically relevant topic based on all levels outlined in Bloom’s Taxonomy ^14^. Specifically, we selected MCQs from the Taiwan Psychiatry Licensing Examination from the past two years (2023 and 2024). These exams represent the first stage of obtaining board certification and consist entirely of MCQs. Questions from these exams are primarily derived from the 12th edition of the Kaplan & Sadock’s Synopsis of Psychiatry ^15^, ensuring that the content reflects the latest psychiatric knowledge. These questions were crafted by experienced board-certified psychiatrists, and each exam consisted of 100 questions, each worth one point. A score of 60 is set as the passing threshold. To maintain relevance and consistency, questions involving Taiwan-specific laws and policies or those solely testing basic medical knowledge were excluded. The remaining questions were categorized into six domains: pathophysiology and epidemiology, diagnostic assessment and clinical examination, psychopharmacology and other therapeutic modalities, psychosocial and cultural influences, neuroscience and behavioral science, and forensic psychiatry and ethic.

### 2.2. Chatbot Selection

We selected chatbots based on their rankings from the LMarena website ^15^, which provides comparative evaluations of multiple chatbots. This platform allows users to input queries and compare responses from two chatbots, with the final rankings determined by aggregated user choices. The top 10 ranked chatbots were initially screened in this study, excluding those with restricted availability (e.g., geofenced in mainland China). This process identified leading chatbot companies for inclusion. The final selection comprised chatbots from OpenAI, Google, Anthropic, Meta, xAI, Alibaba, and Mistral. We included all available models from each company to ensure diversity in capabilities and performance. The chatbots evaluated in this study included OpenAI’s GPT-4o, GPT-4o-mini, GPT-o1-preview, GPT-o1-mini, and GPT-4-Turbo; Google’s Gemini-1.5-Pro, Gemini-1.5-Flash, Gemini-1.5-Flash-8B, Gemini-Exp1121, and LearnLM1.5 Pro; Anthropic’s Claude-2, Claude-2.1, Claude-3-Haiku, Claude-3-Sonnet, Claude-3-Opus, Claude-3.5-Haiku, Claude-3.5-Sonnet, and Claude-3.5-Sonnet-June; Meta’s Llama-3.1-70B, Llama-3.1-405B, Llama-3.1-Nemotron, and Llama-3.2-90B; xAI’s Grok-beta; Alibaba’s Qwen-2 and Qwen-2.5; and Mistral’s Large-2.

### 2.3. Evaluation Procedure

The evaluation was conducted from November 29 to December 1, 2024. Each chatbot underwent zero-shot testing with batches of 10 MCQs. The prompt provided to the chatbots was: “Below are 10 multiple-choice questions with their options. Please provide the answers as numbers only.” Limiting each batch to 10 MCQs ensured that all questions fit within the context length constraints of the chatbots. Each chatbot’s responses were recorded, and the correctness of each answer was documented. For a deeper understanding of chatbot reasoning, we further prompted the chatbots to explain their answers for specific questions using the query: “Please explain your reasoning for Question X in detail.”

The primary outcome was the raw score for each chatbot, which was then converted into a logit ability estimate using the Rasch model. The secondary outcome involved a qualitative analysis of the best-performing chatbot’s strengths and weaknesses, based on its explanations for individual questions. Special attention was given to questions where the chatbot answered difficult questions correctly or simple questions incorrectly.

### 2.4. Statistic Analysis

First, we performed descriptive analysis, including calculations of the mean, standard deviation, maximum and minimum scores, and Cronbach’s alpha to assess the internal consistency of the test. We then conducted further analysis using the Rasch model, a statistical method widely used in psychometrics to evaluate the relationship between item difficulty and the ability of respondents (in this case, chatbots). Rasch analysis is based on modern test theory and provides estimates of item difficulty and respondent ability on the same scale, expressed in logits (log-odds units). The analysis was performed using the WINSTEPS software (version 5.8.2) on a Windows 10 operating system.

In the Rasch model, the probability of a chatbot answering a question correctly is determined by the difference between the chatbot’s ability (β) and the item’s difficulty (δ) ^16^. This is calculated using the formula: P(X = 1) = exp(β - δ) / [1 + exp(β - δ)], where P(X = 1) represents the probability of a correct response. When the chatbot’s ability (β) matches the difficulty of the item (δ), the probability of answering correctly is 50%. The Rasch model iteratively adjusts these estimates to minimize error, producing reliable measures of both ability and difficulty.

Before conducting the Rasch analysis, we first checked whether the dataset met the model’s basic assumptions. This involved using principal component analysis (PCA) on the residuals to ensure that the test mainly measured one underlying concept. In Rasch analysis, it’s important that secondary dimensions (other factors besides the main concept) explain no more than 20% of the residual variance ^4, 17^. We also calculated essential unidimensionality, which shows how much of the variance is explained by the main concept—in this case, psychiatric clinical knowledge. A value of 50% or higher was considered acceptable ^18^. After confirming unidimensionality, we estimated chatbot ability and item difficulty using joint maximum likelihood estimation (JMLE). The passing threshold was set at 60% accuracy for the select-ed questions and was also estimated using JMLE. To evaluate whether the best-performing chatbot significantly surpassed this threshold, we performed Wald tests, considering a two-tailed p-value of < 0.05 as statistically significant.

To ensure the model’s validity, we analyzed fit statistics to determine if the data aligned with the expectations of the Rasch model. Infit mean square (MNSQ) was utilized to evaluate the consistency of responses to items matching the chatbot’s ability level, while outfit MNSQ detected unusual response patterns to extremely easy or difficult items. Acceptable values for both metrics ranged from 0.5 to 1.5 ^19^. Additionally, z-standardized (ZSTD) was evaluated, with acceptable values between ±1.96 ^19^.

Finally, we created a person–item map (PKMAP) for each chatbot, providing a visual representation of its performance across questions of varying difficulty and showing how its ability matches the difficulty levels of the test items. We conducted a detailed analysis of its responses to identify patterns (as mentioned in the evaluation procedure section), with particular focus on the simplest questions it answered incorrectly and the most challenging ones it answered correctly.

## 3. Results

### 3.1. Chatbot Performance Overview

Table 1 listed the distribution of questions across two years of licensing exams, comprising 160 questions divided into six categories. The majority of questions were categorized under Pathophysiology and Epidemiology, Diagnostic Assessment and Clinical Examination, and Psychopharmacology and Other Therapeutic Modalities. Table 2 lists the chatbots analyzed in this study, along with their release dates and associated companies. These chatbots were released between July 2023 and November 2024. We evaluated the performance of 27 chatbots (Table 3), yielding an average raw score of 97.7 (61% accuracy), with a standard deviation of 19.5. The highest score was 129, while the lowest was 56. The test reliability Cronbach’s alpha was 0.93.

**Table 1.**
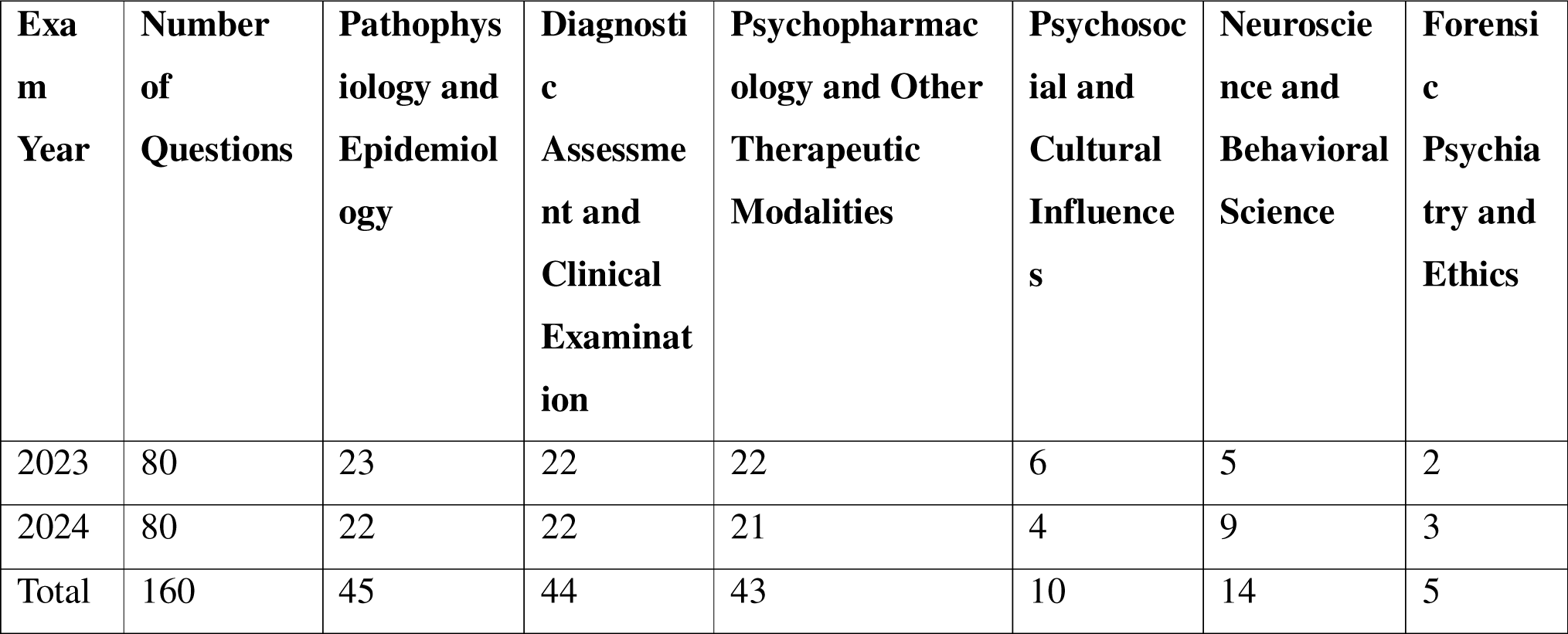
Distribution of Question Characteristics.

| Exam Year | Number of Questions | Pathophysiology and Epidemiology | Diagnostic Assessment and Clinical Examination | Psychopharmacology and Other Therapeutic Modalities | Psychosocial and Cultural Influences | Neuroscience and Behavioral Science | Forensic Psychiatry and Ethics |
| --- | --- | --- | --- | --- | --- | --- | --- |
| 2023 | 80 | 23 | 22 | 22 | 6 | 5 | 2 |
| 2024 | 80 | 22 | 22 | 21 | 4 | 9 | 3 |
| Total | 160 | 45 | 44 | 43 | 10 | 14 | 5 |

**Table 2.** Chatbot Characteristics.

| Model Name | Company | Released Month |
| --- | --- | --- |
| ChatGPT-4o-latest | OpenAI | 2024/11 |
| Gemini-1.5-Pro | Google | 2024/9 |
| Gemini-1.5-Flash | Google | 2024/9 |
| Gemini-1.5-Flash8B | Google | 2024/9 |
| LearnLM1.5 Pro | Google | 2024/11 |
| Claude-3.5-Sonnet | Anthropic | 2024/10 |
| GPT-4o | OpenAI | 2024/05 |
| 4o-mini | OpenAI | 2024/07 |
| o1-preview | OpenAI | 2024/09 |
| o1-mini | OpenAI | 2024/09 |
| Gemini-Exp1121 | Google | 2024/11 |
| Claude-3.5-Sonnet-June | Anthropic | 2024/06 |
| GPT-4-Turbo | OpenAI | 2023/11 |
| Mistral-Large-2 | Mistral | 2024/07 |
| Claude-3-Sonnet | Anthropic | 2024/03 |
| Claude-3-Haiku | Anthropic | 2024/03 |
| Claude-3-Opus | Anthropic | 2024/03 |
| Qwen2-72B | Alibaba | 2024/06 |
| Claude-2 | Anthropic | 2023/07 |
| Claude-2.1 | Anthropic | 2023/11 |
| Qwen-2.5-72B | Alibaba | 2024/09 |
| Claude-3.5-Haiku | Anthropic | 2024/10 |
| Llama-3.2-90B | Meta | 2024/09 |
| Llama-3.1-70B | Meta | 2024/07 |
| Llama-3.1-405B | Meta | 2024/07 |
| Llama-3.1-Nemotron | Meta | 2024/09 |
| Grok-beta | xAI | 2024/11 |

**Table 3.** Raw Scores and Rasch Model Parameters for Chatbots.

| Model Name | Raw score | JMLE measure | JMLE SE | Infit MNSQ | Infit ZSTD | Outfit MNSQ | Outfit ZSTD |
| --- | --- | --- | --- | --- | --- | --- | --- |
| ChatGPT-4o-latest | 117 | 1.47 | 0.24 | 0.8912 | -0.7891 | 0.9468 | -0.0991 |
| Gemini-1.5-Pro | 117 | 1.47 | 0.24 | 0.7862 | -1.6592 | 0.6234 | -1.4494 |
| Gemini-1.5-Flash | 110 | 1.1 | 0.22 | 1.1754 | 1.4112 | 1.1662 | 0.7412 |
| Gemini-1.5-Flash8B | 77 | -0.33 | 0.2 | 1.1408 | 1.6011 | 1.3981 | 1.6614 |
| LearnLM1.5 Pro | 117 | 1.47 | 0.24 | 1.0241 | 0.221 | 0.8744 | -0.3691 |
| Claude-3.5-Sonnet | 116 | 1.41 | 0.23 | 0.6761 | -2.7093 | 0.4657 | -2.3495 |
| GPT-4o | 120 | 1.64 | 0.24 | 0.7869 | -1.5692 | 0.7419 | -0.8193 |
| 4o-mini | 76 | -0.38 | 0.2 | 1.0415 | 0.511 | 0.9184 | -0.2891 |
| o1-preview | 129 | 2.23 | 0.27 | 1.0829 | 0.5411 | 0.9724 | 0.071 |
| o1-mini | 77 | -0.33 | 0.2 | 1.1953 | 2.1812 | 2.4051 | 4.6324 |
| Gemini-Exp1121 | 116 | 1.41 | 0.23 | 0.7307 | -2.1893 | 0.4838 | -2.2395 |
| Claude-3.5-Sonnet-June | 115 | 1.36 | 0.23 | 0.7599 | -1.9592 | 0.687 | -1.2193 |
| GPT-4-Turbo | 107 | 0.95 | 0.22 | 1.0189 | 0.201 | 1.0926 | 0.4711 |
| Mistral-Large-2 | 56 | -1.2 | 0.21 | 1.3743 | 3.5814 | 3.0949 | 4.2431 |
| Claude-3-Sonnet | 89 | 0.15 | 0.2 | 0.9344 | -0.6891 | 0.9041 | -0.4291 |
| Claude-3-Haiku | 79 | -0.25 | 0.2 | 1.2249 | 2.4712 | 1.7242 | 2.8217 |
| Claude-3-Opus | 106 | 0.9 | 0.22 | 0.8854 | -0.9991 | 1.0068 | 0.101 |
| Qwen2-72B | 111 | 1.15 | 0.23 | 1.1809 | 1.4312 | 1.3014 | 1.2013 |
| Claude-2 | 67 | -0.74 | 0.2 | 0.9869 | -0.119 | 0.991 | 0.051 |
| Claude-2.1 | 71 | -0.58 | 0.2 | 1.0733 | 0.8711 | 1.1703 | 0.7412 |
| Qwen-2.5-72B | 110 | 1.1 | 0.22 | 0.7036 | -2.6893 | 0.5247 | -2.3395 |
| Claude-3.5-Haiku | 96 | 0.45 | 0.21 | 0.8675 | -1.3491 | 0.6945 | -1.6493 |
| Llama-3.2-90B | 77 | -0.33 | 0.2 | 1.1292 | 1.4811 | 1.1476 | 0.7011 |
| Llama-3.1-70B | 78 | -0.29 | 0.2 | 1.2029 | 2.2512 | 1.3116 | 1.3613 |
| Llama-3.1-405B | 102 | 0.72 | 0.21 | 0.9241 | -0.6791 | 0.775 | -1.0892 |
| Llama-3.1-Nemotron | 94 | 0.36 | 0.21 | 0.9505 | -0.479 | 0.8934 | -0.4891 |
| Grok-beta | 108 | 1 | 0.22 | 0.7748 | -2.0392 | 0.5969 | -1.9794 |

JMLE, Joint maximum likelihood estimation; SE, standard error; MNSQ, mean square; ZSTD: z-standardized.

### 3.2. Dimensionality and Rasch Model Analysis

The dimensionality analysis confirmed that the dataset aligned with the assumptions of the Rasch model. The raw variance explained by the measures was 38.9%, while the essential unidimensionality calculated as the proportion of Rasch-common variance, reached 76.5%. These values exceed the commonly accepted thresholds of 20% for unidimensionality and 50% for essential unidimensionality, supporting the interpretation that the observed response patterns primarily reflect a single latent trait as psychiatric clinical knowledge.

The Rasch model parameters for chatbots were presented in Table 3. ChatGPT-o1-preview achieved the highest performance among all models, with a JMLE ability score of 2.23, substantially surpassing the passing threshold (JMLE = 0.44, p< .001). Its infit (MNSQ = 1.08, ZSTD = 0.54) and outfit (MNSQ = 0.97, ZSTD = 0.07) statistics were within the acceptable range of 0.5–1.5 and ±1.96, respectively, indicating strong consistency with the Rasch model’s expectations. The chatbot’s responses demonstrated both reliability and validity in assessing psychiatric clinical knowledge.

### 3.3. Performance and reasoning of ChatGPT-o1-preview

The person–item map (PKMAP) for ChatGPT-o1-preview (Figure 1) visually represents its performance across questions of varying difficulty levels. In the PKMAP, the vertical axis represents the difficulty of questions in logits, with more difficult items located higher on the map, while the horizontal axis separates correct responses (on the left) from incorrect responses (on the right). The upper-left quadrant (e.g., Items 52, 54, 79, and 140) of the map highlights areas where ChatGPT-o1-preview demonstrated strong capabilities, successfully answering challenging questions. Conversely, the lower-right quadrant (e.g., Items 27, 36, 42, 43, 77, 92, 106, 117, 131, and 146) indicates areas where the chatbot struggled, with incorrect answers to relatively easier questions. This distribution provides a clear visualization of the chatbot’s strengths and weaknesses in its performance on the questions.

**Figure 1.**
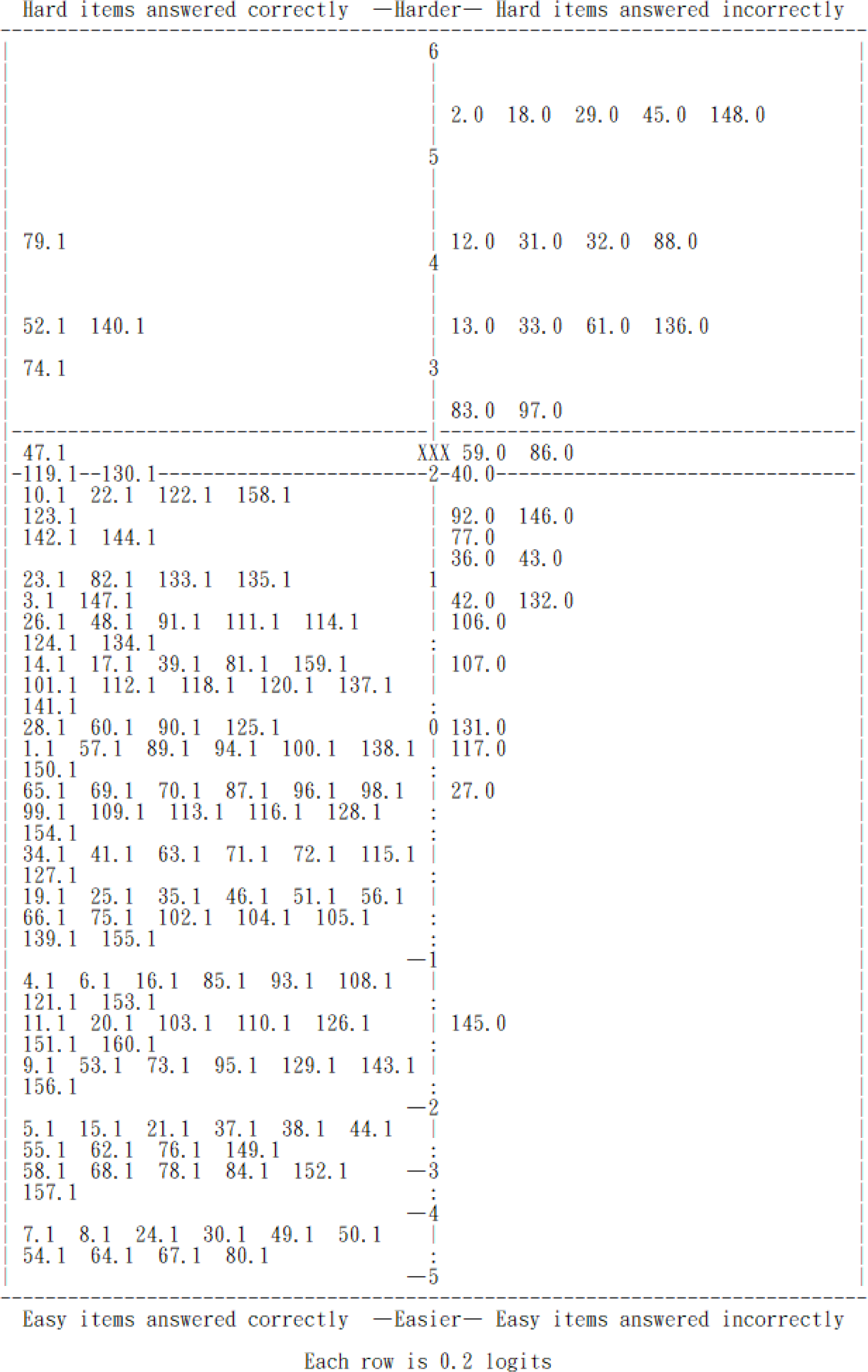
The person–item map (PKMAP) of ChatGPT-o1-preview. It illustrated the relationship between the chatbot’s ability and the difficulty of the test items. The vertical axis, measured in logits, represents the difficulty level of the questions. The symbol “XXX” indicates the chatbot’s estimated ability, and each item on the map represents a specific question number from the exam, accompanied by either a “1” for correct responses or a “0” for incorrect ones. A “1” indicates a correct response and is displayed on the left side of the map, while a “0” signifies an incorrect response and is placed on the right side. The vertical positioning of each item reflects its difficulty.

A detailed analysis of the chatbot’s answering process (Table 4, Table 5) revealed key strengths and weaknesses of its reasoning. ChatGPT-o1-preview excelled in areas such as diagnostic reasoning and broader therapeutic concepts (e.g., recognizing paraphilic disorders and treatment paradigms for schizophrenia), and pharmacological principles (e.g., drug mechanisms, indications, and side effects). However, it exhibited notable limitations in recalling specific factual details (e.g., remission timelines for transvestic disorder, concordance rates for generalized anxiety disorder in twin studies, or DSM-5-TR definitions of negative symptoms). Additionally, biases in reasoning were observed, such as overemphasizing lithium’s efficacy in depression augmentation therapy while underestimating the role of antipsychotics or dismissing hypnosis as a therapeutic option. The chatbot also demonstrated its capacity for self-correction. In several cases (e.g., Items 27, 42, 77, 131, and 145), it revised its initial incorrect answers upon re-evaluation, ultimately producing accurate solutions.

**Table 4.**
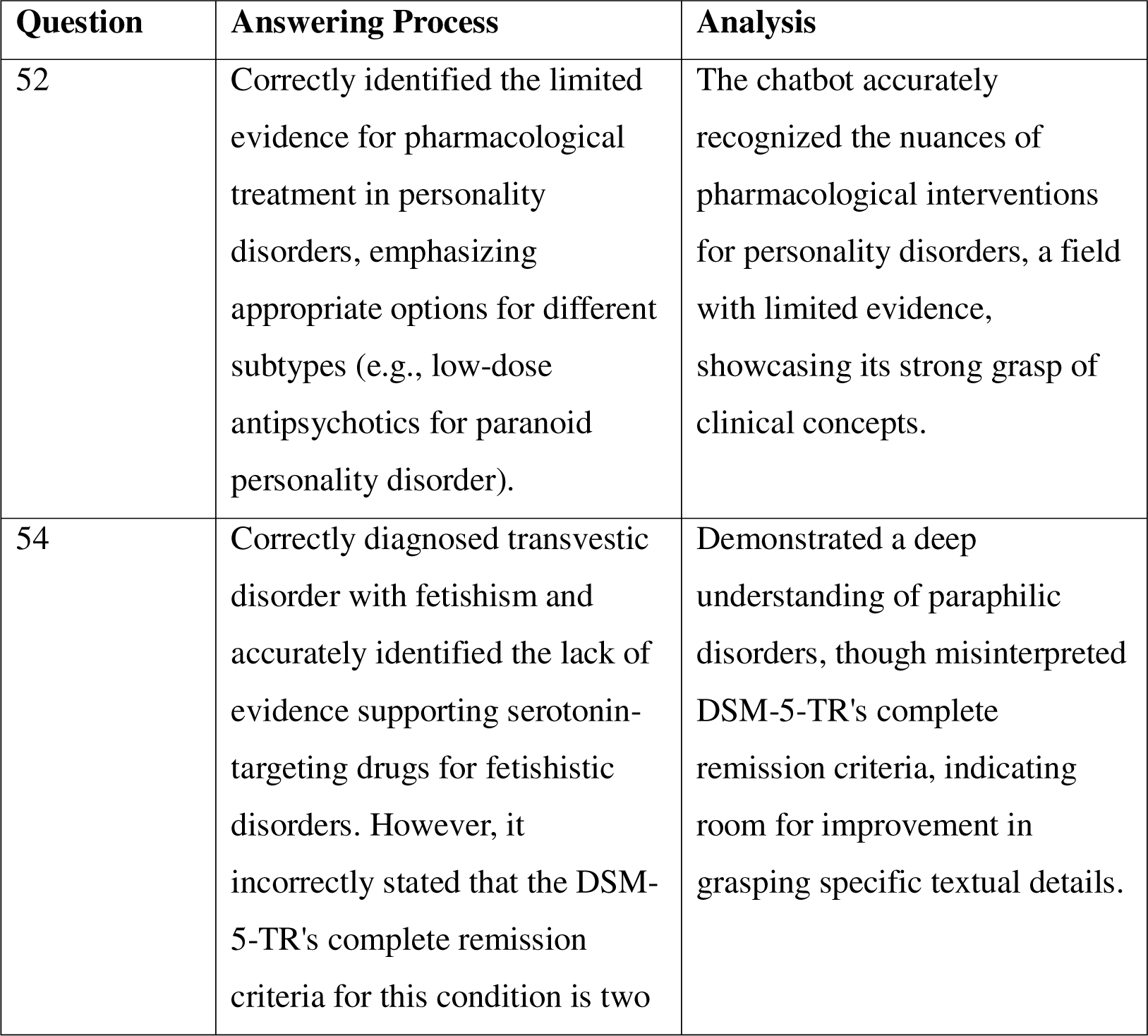
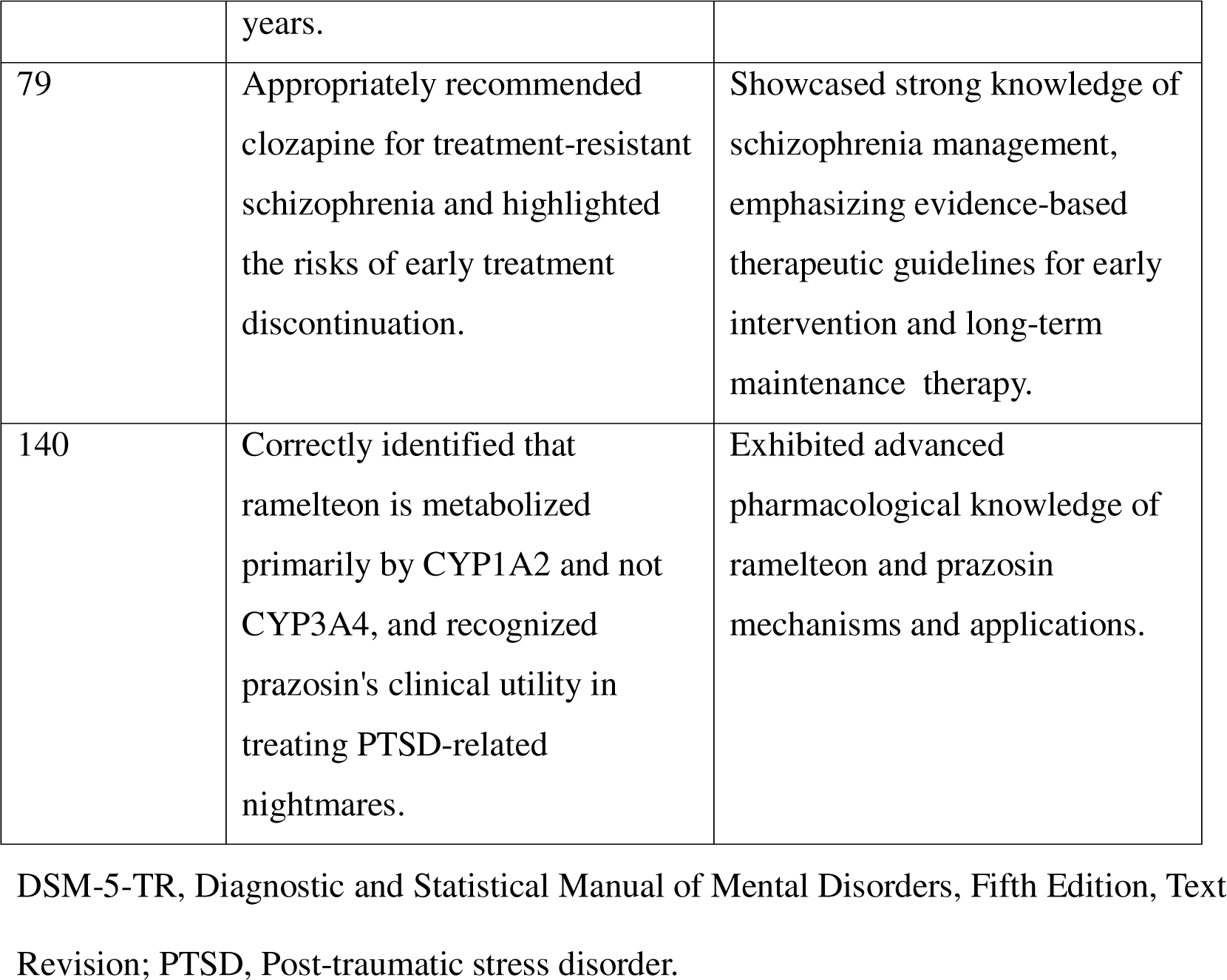
Strengths Analysis of ChatGPT-o1-preview’s Answering Process Question Answering Process Analysis.

**Table 5.**
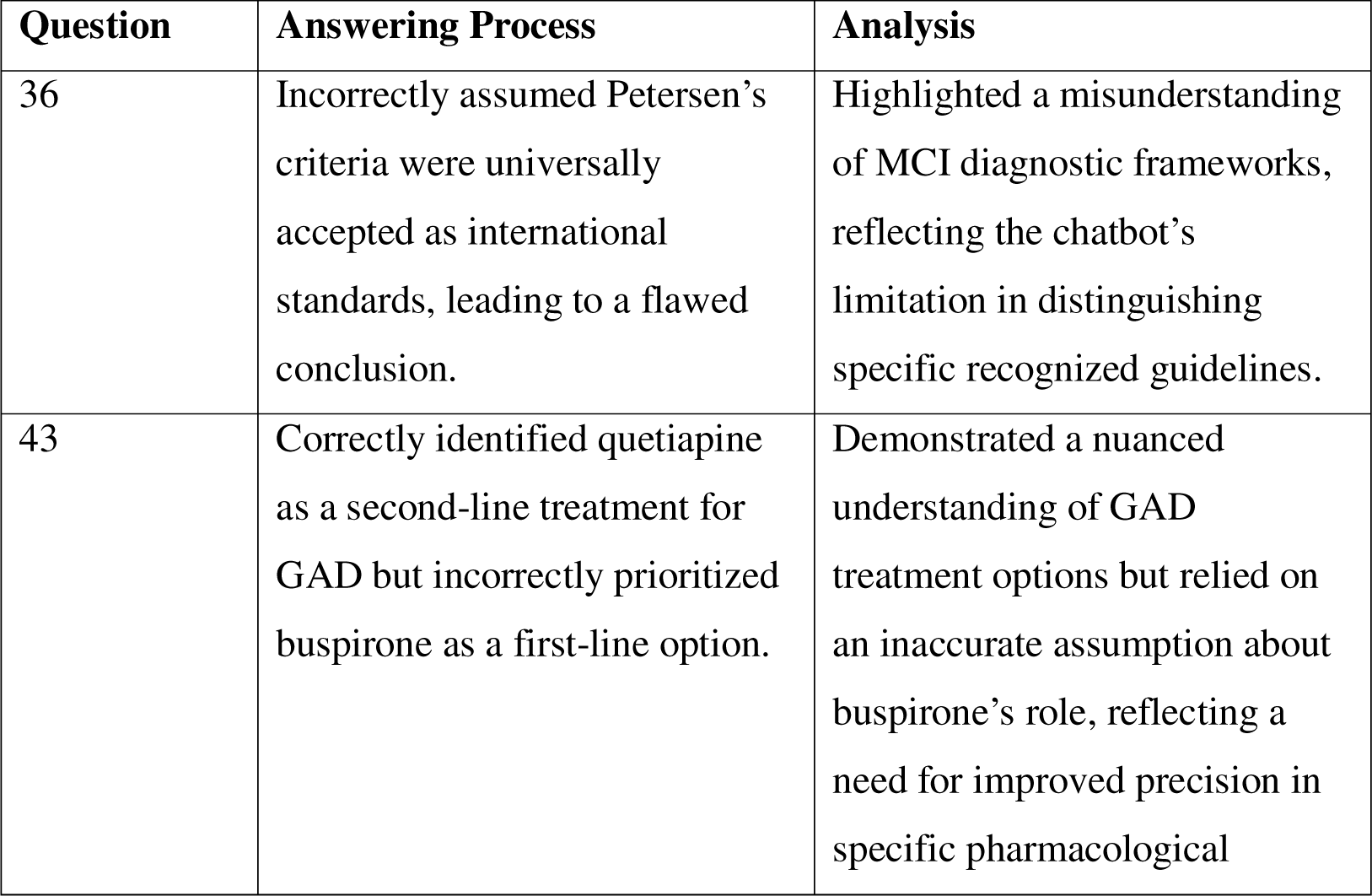
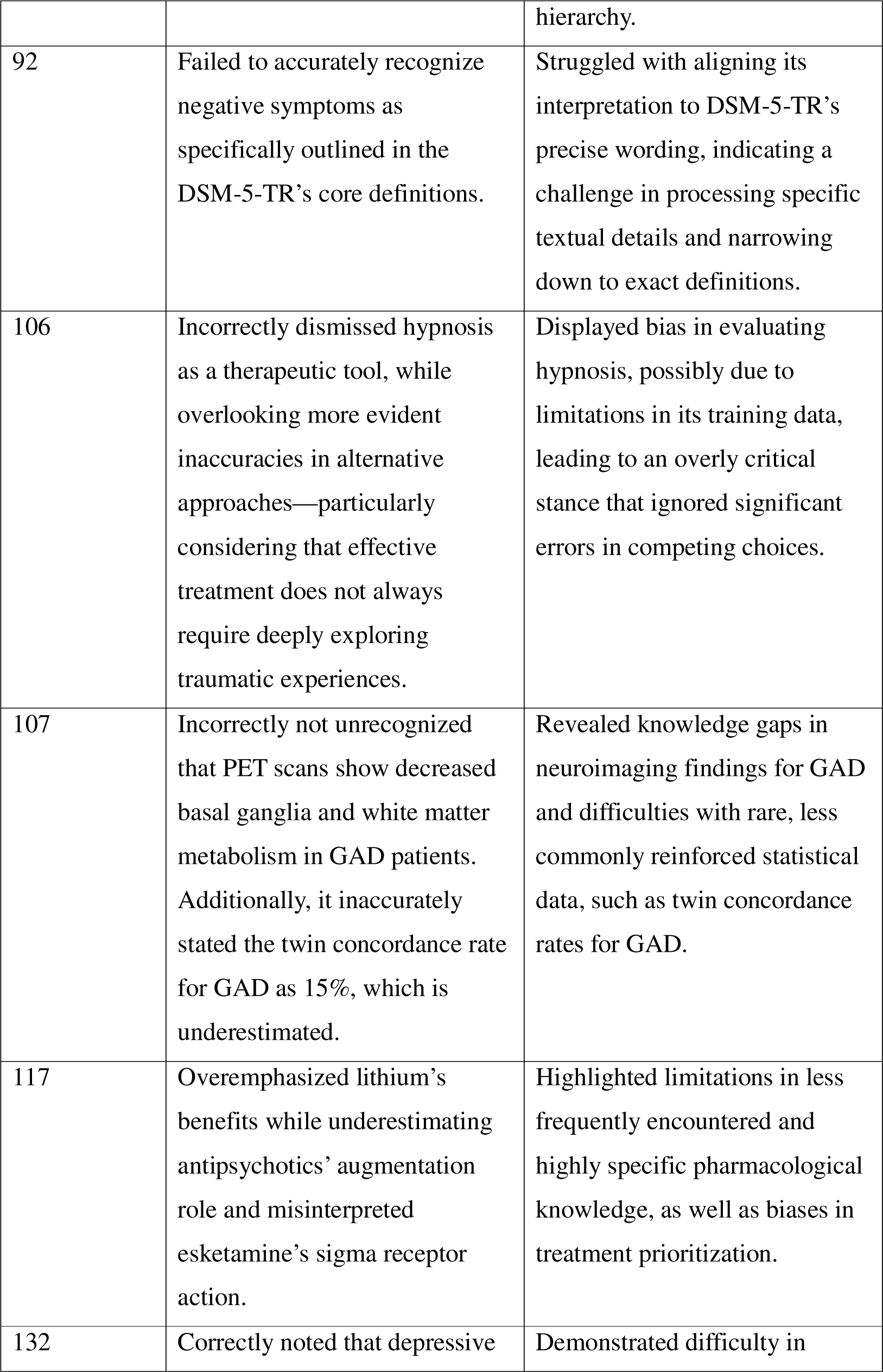
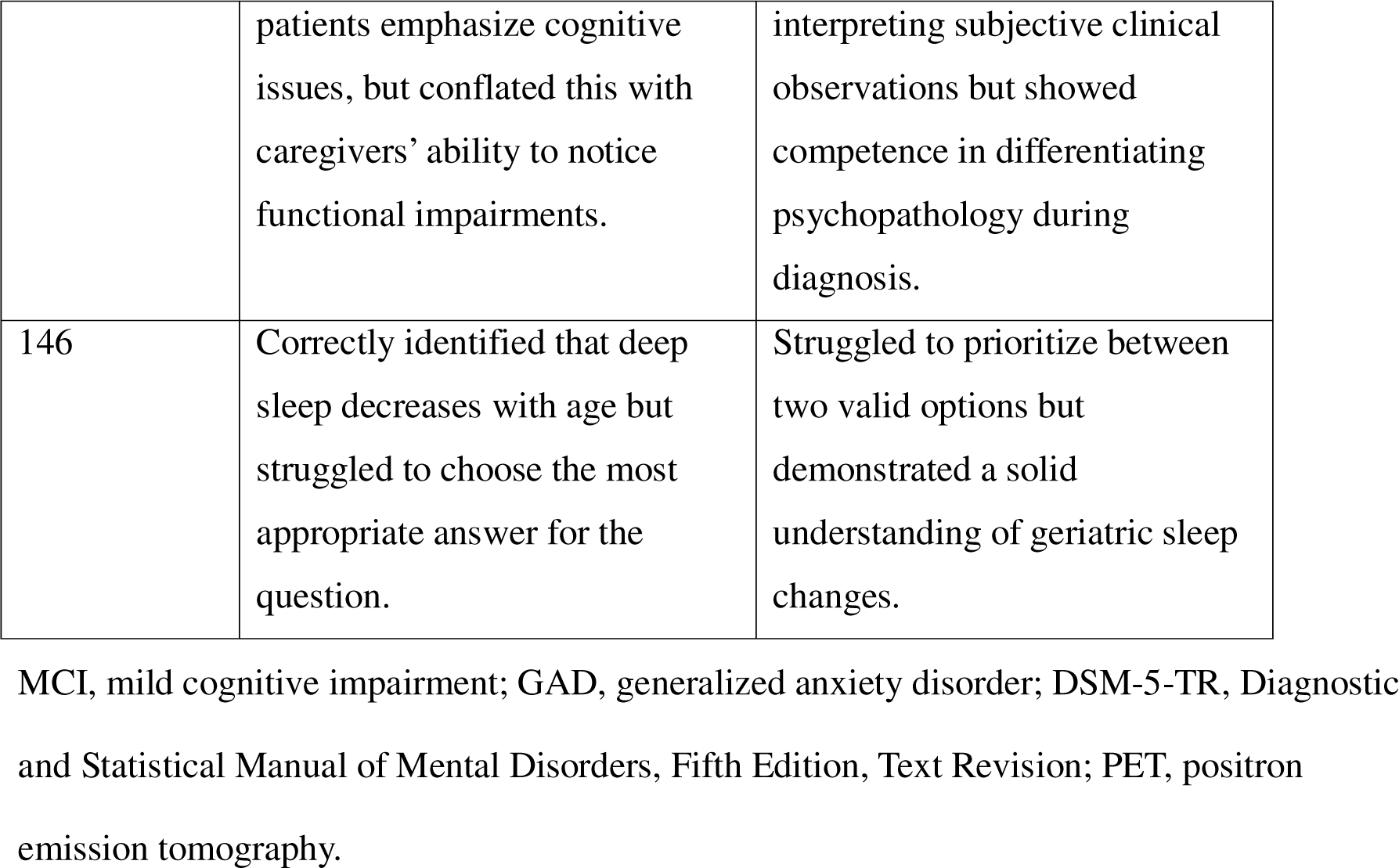
Weaknesses Analysis of ChatGPT-o1-preview’s Answering Process.

## 4. Discussion

To our knowledge, this study is the first to apply Rasch analysis to evaluate chatbots’ clinical knowledge in psychiatry using expert-designed multiple-choice questions. Among the 27 chatbots assessed, ChatGPT-o1-preview emerged as the top-performing model, achieving a correct response rate of 80.6% and a JMLE ability estimate of 2.23. According to the Rasch model, this indicates that ChatGPT-o1-preview would achieve a correct response rate of 85.6% on questions with difficulty at the passing threshold. The strengths of ChatGPT-o1-preview were particularly evident in its understanding of high-level diagnostic, therapeutic, and pharmacological concepts. For instance, the model showcased advanced reasoning in areas such as schizophrenia management across various stages, diagnostic clarity in paraphilic disorders, and a thorough understanding of psychopharmacology, including drug mechanisms, indications, and side effects. These strengths reflect the depth of training data and the reasoning capabilities of the underlying large language model (LLM). Moreover, ChatGPT-o1-preview exhibited a strong ability to self-correct during re-evaluation.

Despite these strengths, the chatbot demonstrated notable limitations. It struggled with questions requiring precise factual recall, such as the exact Diagnostic and Statistical Manual of Mental Disorders, Fifth Edition, Text Revision (DSM-5-TR) criteria ^20^, remission timelines for transvestic disorder, and rare statistical data (e.g., concordance rates for generalized anxiety disorder). This highlights the need for caution regarding the chatbot’s susceptibility to hallucinations or confalubations ^21^. When probed for highly specific details, ChatGPT-o1-preview occasionally generated inaccurate or fabricated information. This limitation is consistent with prior studies on chatbot performance in factual recall tasks. For example, the SimpleQA benchmark ^22^, which focuses on short, fact-seeking queries from diverse aspects, requires each question to meet strict criteria: it must have a single, indisputable answer that is easy to grade, and the answer should remain constant over time. However, chatbots tend to perform poorly on SimpleQA tasks. Similarly, the Chinese SimpleQA ^23^, a localized version of the SimpleQA benchmark, has demonstrated similar challenges in chatbot performance. As a result, while chatbots can provide valuable insights, their outputs must be carefully verified, particularly when addressing specific clinical details.

Additionally, the chatbot occasionally displayed reasoning biases, such as overemphasizing lithium’s efficacy in depression augmentation therapy while underestimating the role of antipsychotics or dismissing hypnosis as a viable treatment for dissociative identity disorder. Such biases may stem from the training data’s inherent limitations or uneven exposure to certain clinical concepts ^24-26^. Such findings align with previous research, including biasmedQA, which highlights how biased information in prompts can lead to biased clinical judgments by chatbots ^27^.

Our study highlights the influence of training data availability on chatbot performance. While ChatGPT-o1-preview demonstrated superior knowledge in broadly discussed psychiatric topics, its performance in niche areas was less reliable. This discrepancy reflects the accessibility of training resources; open-access journals, books, and widely available materials likely dominate the training corpus, while proprietary textbooks and leading psychiatric journals remain less accessible. Recent partnerships between publishers and chatbot developers signal potential solutions to this issue. For instance, Axel Springer has partnered with OpenAI to integrate journalism with AI technologies ^28^, and Elsevier Health has collaborated with OpenEvidence to develop ClinicalKey AI ^29^, a tool designed to enhance medical decision-making using artificial intelligence.

Collaborative efforts such as these could bridge the gap between accessible and proprietary knowledge, benefiting chatbot training. Techniques like retrieval-augmented generation (RAG)^30, 31^, which uses vectorized data to enhance responses by retrieving relevant information, or fine-tuning ^32, 33^, which optimizes models by training them on domain-specific datasets, can further improve chatbot accuracy in specialized fields. By leveraging such advancements, chatbots could become more reliable tools for addressing complex and niche clinical scenarios.

From a clinical perspective, the findings suggest that chatbots like ChatGPT-o1-preview show significant potential as tools for delivering timely and reasonably accurate feedback to support clinical decision-making. However, clinicians should use these tools with caution. First, given the chatbot’s performance limitations, it is advisable to present the chatbot with pre-defined clinical options and prompt it for explanations, rather than relying on it to generate responses independently. With adequate prompt engineering ^34, 35^, which involves designing specific and structured queries, can improve the quality of chatbot responses. Second, encouraging chatbots to perform iterative reasoning or re-evaluate their responses can enhance accuracy, as demonstrated in this study.

When incorporating chatbots into clinical workflows, consulting multiple chatbots simultaneously can provide diverse perspectives, helping to mitigate the biases inherent in individual models. By framing queries that include the clinician’s own hypotheses or reasoning, chatbots can be guided to provide more focused responses. It is also prudent to avoid relying on chatbots for factual recall and instead use them for reasoning and decision support. Comparing the outputs of different models can help identify consistent patterns and discrepancies, which can then be synthesized to inform clinical judgment. Ultimately, chatbot-generated responses should serve as references rather than definitive conclusions, ensuring that clinicians retain responsibility for decision-making. Through thoughtful integration, chatbots can become valuable adjuncts to clinical expertise while minimizing risks and enhancing patient care.

The rapid advancements in chatbot capabilities cannot be ignored. Historical research suggests that as of early 2024, chatbots had achieved performance sufficient to pass licensing exams, with some models, such as ChatGPT-o1-preview, significantly surpassing that benchmark ^4^. This accelerated improvement suggests that conclusions from earlier studies, particularly those conducted in 2023 or earlier ^36, 37^, may no longer accurately reflect the current state of chatbot capabilities. Therefore, ongoing evaluation of new chatbot iterations is essential to keep pace with their evolving capabilities and to better understand their applicability in clinical practice.

This study has several limitations. First, the sample size of 27 chatbots, although sufficient for a pilot study using Rasch analysis ^38^, may limit the generalizability of our findings. However, the large number of questions used (n = 160) ensured a low standard error for ability estimates, enhancing the reliability of the results. Second, while the multiple-choice questions used in this study provided a standardized method of assessing psychiatric knowledge, they may not fully capture the breadth of clinical expertise required in practice. Third, questions specific to regional laws and policies were excluded, which may limit the applicability of the findings to regions with differing regulations. Fourth, all questions were presented in traditional Mandarin, and the results may vary across different languages. Lastly, while possessing clinical knowledge is important, it does not necessarily translate to the effective application of that knowledge in real-world scenarios. Although we have highlighted key aspects of a potential clinical workflow, future research should prioritize evaluating the efficacy of chatbots in supporting clinical decision-making through controlled trials in psychiatric settings ^7, 39^.

## 5. Conclusion

This study demonstrates that ChatGPT-o1-preview, released by OpenAI in September 2024, outperformed other chatbots in a standardized evaluation of psychiatric clinical knowledge. The model’s strengths lie in its understanding of diagnostic frameworks, treatment paradigms, and pharmacology concepts. However, its limitations in recalling specific details, addressing niche knowledge, and overcoming biases highlight the need for cautious interpretation of its outputs. Building on these findings, we have proposed key aspects of a potential clinical workflow to guide the practical integration of chatbots into psychiatric practice. As chatbot technologies continue to evolve, ongoing assessments of their capabilities and controlled clinical trials will be crucial for understanding their broader applicability and ensuring safe and effective implementation in clinical settings.

## Data Availability

All data produced in the present study are available upon reasonable request to the authors.

## Notes

### Competing Interest Statement

The authors have declared no competing interest.

### Funding Statement

This study did not receive any funding.

